# S4-Multi: enhancing polygenic score prediction in ancestrally diverse populations

**DOI:** 10.1101/2025.01.24.25321098

**Authors:** John Baierl, Jonathan P. Tyrer, Ping-Hung Lai, Simon A. Gayther, Yi-Wen Hsiao, Michelle Jones, Pei-Chen Peng, Paul D. P. Pharoah

## Abstract

While polygenic scores (PGSs) have shown promise in advancing precision medicine by capturing the additive effects of common germline variants on inherited disease risk, they are presently limited by reduced performance outside of European-origin populations. We extend our previously developed Bayesian polygenic model (PGM) method, select and shrink with summary statistics (S4), to improve prediction accuracy in ancestrally diverse populations. We benchmark this multi-ancestry extension (S4-Multi) against alternative methods on both simulated and biobank data predicting type 2 diabetes, breast cancer, colorectal cancer, asthma, and stroke. In simulation tests, we find that S4-Multi achieves 169% improvement on average over its single ancestry S4 counterpart at prediction in non-European target populations. S4-Multi matches or exceeds top performing methods across the ancestry continuum. In biobank tests, we find that the top-performing PGM method varies considerably by target ancestry and phenotype, with S4-Multi achieving comparable performance to top multi-ancestry methods overall. However, S4-Multi does so while including between 9% and 77% fewer genetic variants relative to competing models, suggesting potential for robust performance in clinical settings with limited available genomic data.

## Introduction

The heritable component of most disease risk is polygenic in nature^1,2^, and polygenic scores (PGSs) have shown promise in refining genetic risk prediction and personalized medicine in recent years^3–6^. PGSs are able to capture the additive effects of common germline genetic variants on disease risks, including those that fail to reach genome-wide significance on their own, producing a single weighted sum for each individual that is more strongly associated with the phenotype of interest. PGSs have proven useful estimating the probabilistic susceptibility of individuals to a range of complex traits, showing validity in both research-based case-control studies and population-based cohort studies^7–9^. Genetic risk estimation holds the potential to aid in early disease detection, improve the effectiveness of screening programs, and inform preventative intervention choices both used alone and in conjunction with other lifestyle and clinical risk factors^10–12^. PGSs are currently incorporated in the comprehensive breast and ovarian cancer risk models implemented in the CanRisk Tool, providing guidance for health care professionals in the patient consultation process^13,14^.

A PGS is computed by applying a polygenic model (PGM), which consists of a set of variants and their associated weights, to an individual’s genotypes at those variants. Multiple methods have been developed for PGM construction. These differ in their approaches for selecting variants for inclusion in the model and how those variants are weighted. Hard threshold approaches like linkage disequilibrium (LD) clumping and thresholding (CT) select SNPs based on a single *p*-value threshold. LD information is then accounted for by removing highly correlated SNPs, with weights corresponding to the SNP-specific effect sizes. Other methods employ regularization on a much larger set of variant effect sizes. These allow for multiple correlated SNPs at each locus to be included in the model by accounting for local LD structure between loci. Examples include LDpred2, polygenic risk score-continuous shrinkage (PRS-CS), and normal-mixture models^15–17^. These computationally intensive PGM methods have been shown to outperform simpler approaches across a range of settings.

Despite the successes of PGMs, several barriers limit their present portability to clinical use. One of the most challenging obstacles is the reduced accuracy of PGMs in non-European (EUR)-origin populations^18–20^. A major reason for this is the continuing overrepresentation of European populations in genome-wide association studies (GWAS) to date^21^. According to the GWAS Diversity Monitor, as of February 2024, 95% of GWAS participants were of European ancestry, while only 3.7% and 0.2% of participants were of Asian and African ancestries, respectively^22^. Cross-ancestry PGM prediction performance has been shown to decrease as the genetic distance between training and validation populations grows, resulting in poor generalizability to non-EUR origin populations^23^. The underlying genetic architecture of disease can also vary by population, further amplifying these challenges^24,25^. PGS prediction in individuals with African (AFR) ancestries has proven particularly challenging due to the higher degree of genetic diversity in those populations^18,26,27^. As a result, current PGM methods risk exacerbating existing health disparities by systematically offering more accurate risk stratification for populations of European descent.

The equitable clinical application of PGMs depends on the development of methods for generalizing across populations. Multi-ancestry PGM methods that integrate data from across the ancestry continuum and incorporate ancestry-specific local LD structure have shown improved predictive performance over those that only utilize GWAS data within a single group of ancestries, including models developed only on training data matching the target ancestries^28–30^. Several PGM methods have been developed in recent years to improve PGS performance in cross-ancestry prediction tasks. PRS-CSx and LDpred2 are two such examples that have demonstrated improved performance in cross-ancestry prediction tasks over common single-ancestry methods^16,31^.

In this paper, we present a multi-ancestry extension of the “select and shrink with summary statistics” (S4) method, a Bayesian PGM that employs continuous shrinkage priors on effect sizes^32^. S4 extends the regularization properties of PRS-CSx by imposing additional penalization of rarer variants. This has previously been shown to achieve prediction accuracy matching or exceeding other computationally intensive PGM methods at single-ancestry prediction^33^. We compare the cross-ancestry predictive performance of the multi-ancestry S4 (S4-Multi) with other commonly used alternatives across a range of complex traits with different genetic architectures. This comparison uses both simulated genetic data developed for benchmarking^34^ and large-scale biobank data from the UK Biobank (UKB), FinnGen, Biobank Japan (BBJ), *All of Us* (AoU), and Global Biobank (GBB). We replicated the simulation and testing structure used by Zhang et al 2023^34^, a recent and comprehensive comparison of twelve PGS methods on multi-ancestry prediction tasks, adding three S4-based methods for model comparison and benchmarking.

## Methods

### S4 Overview

The single-ancestry S4 method was previously presented in Dareng et al 2022^32^. We review the main ideas here to highlight the extensions to the multi-ancestry case. Generally, a polygenic model comprises a set of variants associated with the phenotype together with their weights. The PGS for an individual is an application of a polygenic model to the genotype of the individual (*j*) and is the sum of the allele dosage at each variant (*i*) (*x*_*ij*_ ∈ {0,1,2}) weighted by its corresponding effect size (*w*_*i*_) over the *i* variants included in the model. So, the PGS for the *j*^th^ individual (*PGS_j_*) is given by the linear function:

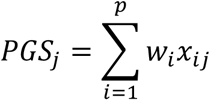

The PGM weights correspond to the log odds ratios from a case-control study for a disease trait. For a quantitative trait, the weights correspond to the beta coefficients from a linear regression model. The central tasks in any PGM construction consist of: (1) selecting which SNPs to include in the model, and (2) adjusting their weights to impose shrinkage or other desired model properties.

In the selection stage of S4 (*Fig. 1*, development stage 1), variants from a GWAS are ranked and selected based on their statistical significance and correlation with previously selected SNPs. S4 partitions SNPs into roughly independent genomic regions or blocks, performing SNP selection separately on each block to reduce computational cost. Top-ranked SNPs are iteratively added if their correlation with all other included SNPs (*r*^2^) is less than 0.85. New SNPs are put into groups with which they are best correlated, with a new group started when a newly added SNP has a correlation below 0.02 with other selected SNPs. The selection procedure terminates when the *p*-value/*r*^2^ ratio for candidate SNPs reaches a pre-determined threshold.

**Fig. 1:**
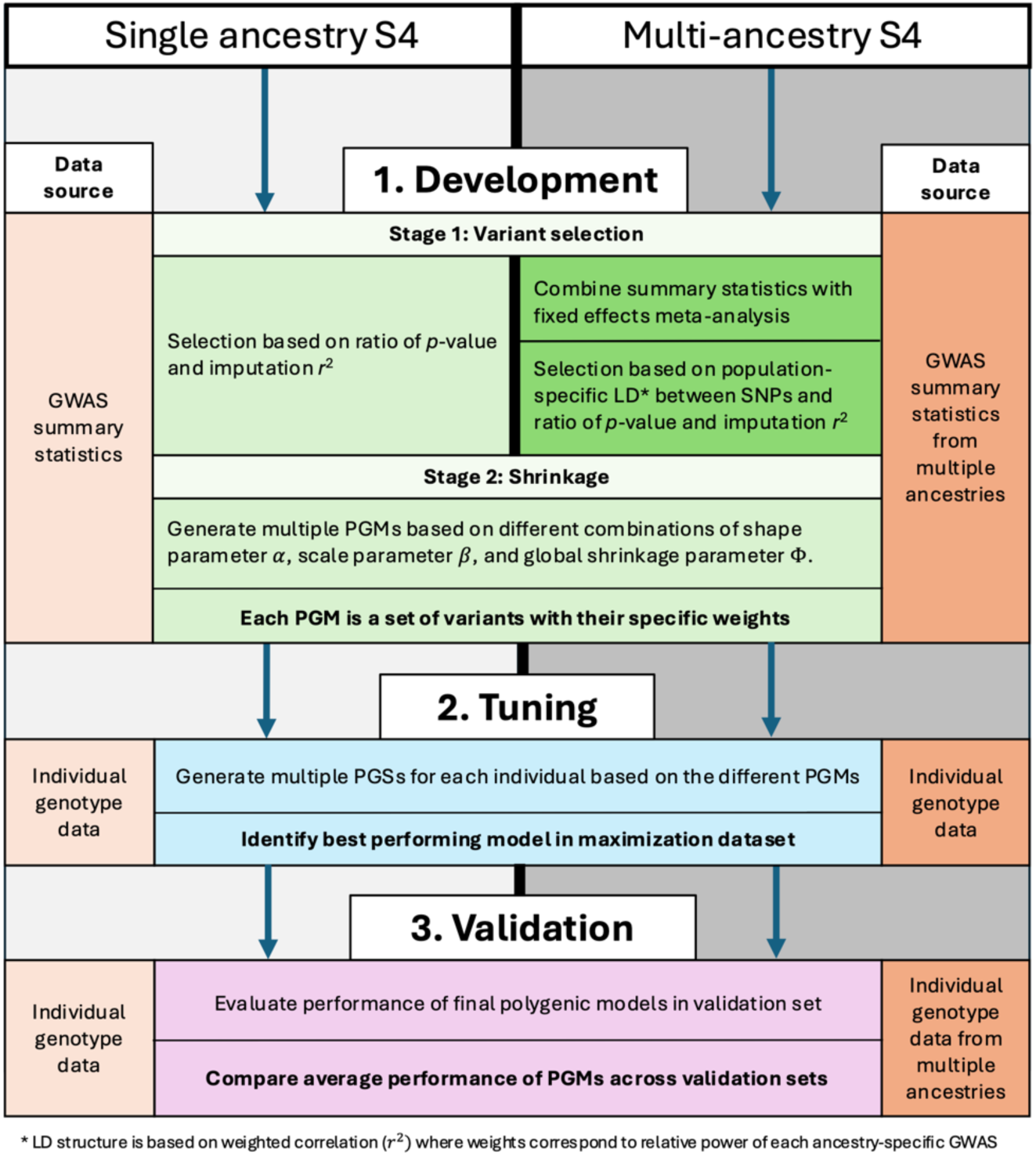
Comparison of pipelines for single and multi-ancestry S4 development, tuning, and validation

S4 adjusts SNP weights (*Fig. 1*, development stage 2) in a similar manner to PRS-CSx, employing a Bayesian model construction that places a shared global-local continuous shrinkage prior on effect sizes to impose sparsity^31^. The SNP-specific shrinkage parameter, Ψ_*j*_, is assigned a gamma-gamma prior with two additional hyperparameters, *α* and *β*, that control the shrinkage of effect sizes around 0 and shrinkage of larger effect sizes, respectively. So, the full hierarchical prior specification is given by:

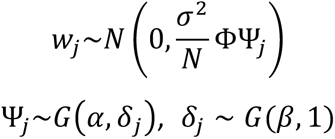

with residual variance *σ*^2^, sample size *N*, and a global shrinkage parameter Φ shared across genetic markers. However, S4 extends the PRS-CSx algorithm by further adjusting the global shrinkage parameter Φ by dividing by the standard deviation of the summary statistic. Since effect size estimates for rarer variants will tend to carry a greater variance, this effectively imposes a further shrinkage penalty for rarer variants. Multiple polygenic models are then generated over a grid of values for *α*, *β*, and Φ*.

Model tuning requires a second, independent data set to reduce over-fitting. Tuning involves evaluating the multiple polygenic models on individual genomic data. The best performing PGM is then selected for model validation and comparison with models developed using other methods.

### Multi-ancestry Extension

The multi-ancestry implementation of S4 makes several adjustments to account for differences in population structure between the training, tuning, and target populations (*Fig. 1*). At the initial stage, S4-Multi incorporates GWAS summary statistics from multiple ancestries to account for between-population genetic diversity. Ancestry-specific GWAS effect sizes are combined via a fixed-effects meta-analysis to produce SNP-specific weights for subsequent selection and effect-size shrinkage. Population-specific LD patterns are accounted for in the variant-selection stage by computing a weighted average of the correlations from each ancestry-specific GWAS. This is the used for SNP selection in the same manner as the single-ancestry S4 construction.

## Data and Results

We evaluated the prediction accuracy of S4-Multi in two settings. First, we benchmarked S4-Multi against twelve other PGM constructions using simulated genomic data. Then we evaluated S4-Multi against other top-performing PGS methods for predicting a set of complex traits using available GWAS summary statistics and individual genomic data from UK Biobank, FinnGen, Biobank Japan, *All of Us*, and the Global Biobank Initiative.

### Simulated data testing

We replicated the data simulation and testing structure used by Zhang et al^34^, a recent and comprehensive comparison of twelve PGM methods on multi-ancestry prediction tasks. We added three S4-based methods: (1) single-ancestry S4 generated on EUR-only GWAS data; (2) S4-Multi at a *p*-value cutoff of 0.02; and (3) S4-Multi at a *p*-value cutoff of 0.15. Varying the *p*-value threshold allowed for assessing the impact of including more or fewer SNPs in the PGM, comparing both against the single-ancestry S4. The twelve other PGM methods for benchmarking included two single-ancestry methods (CT^35^, LDpred2^16^), two EUR-only methods (best EUR CT, best EUR LDpred2), three weighted PRS methods (weighted CT, weighted LDpred2, PolyPred-S+^36^), three Bayesian methods (XPASS^37^, PRS-CSx, PRS-CSx [all ancestries]^31^), and two model-free superlearning-based methods (CT-SLEB, CT-SLEB [all ancestries])^34^. All methods were run as recommended except for LDpred2, which was applied with minor adjustments that simplified implementation while improving performance. Rather than calculating separate PGSs for each ancestry and then combining results via regression, we estimated LD patterns by weighting SNPs according to their effect sizes in each ancestry before calculating their correlation.

Simulated multi-ancestry genotype data sets were generated under a range of genetic architectures, mimicking the LD structure of Americans (AMR), Africans (AFR), East Asians (EAS), and South Asians (SAS) based on a reference panel from the 1000 Genomes Project^38^. Phenotypes were determined by randomly selecting causal SNPs across the genome, with casual SNP proportions set to either 0.01, 0.001, or 5 × 10^’(^ to compare performance across polygenicity settings. Different heritability and selection patterns were also compared.

Heritability distribution was set to either constant common SNP heritability or constant per-SNP heritability. Selection patterns were set to either strong, mild, or no negative selection pressure. See *Table S1* and Zhang et al for additional details on the data simulation procedure^34^.

Overall results from simulation studies are presented in *Fig. 2* with model performance (prediction *R*^2^) averaged over all tested genetic architectures, training sample sizes, and causal SNP proportions at each ancestry (60 total runs per ancestry). Results further stratified by causal SNP proportion and sample size are provided in *Figs. S1-S3*. Multi-ancestry methods consistently outperformed their single-ancestry counterparts, matching previous results^34^, with S4-Multi achieving 162% and 169% increases in accuracy over the EUR-only S4 implementation at *p*-value cutoffs of 0.02 and 0.15, respectively.

**Figure 2:**
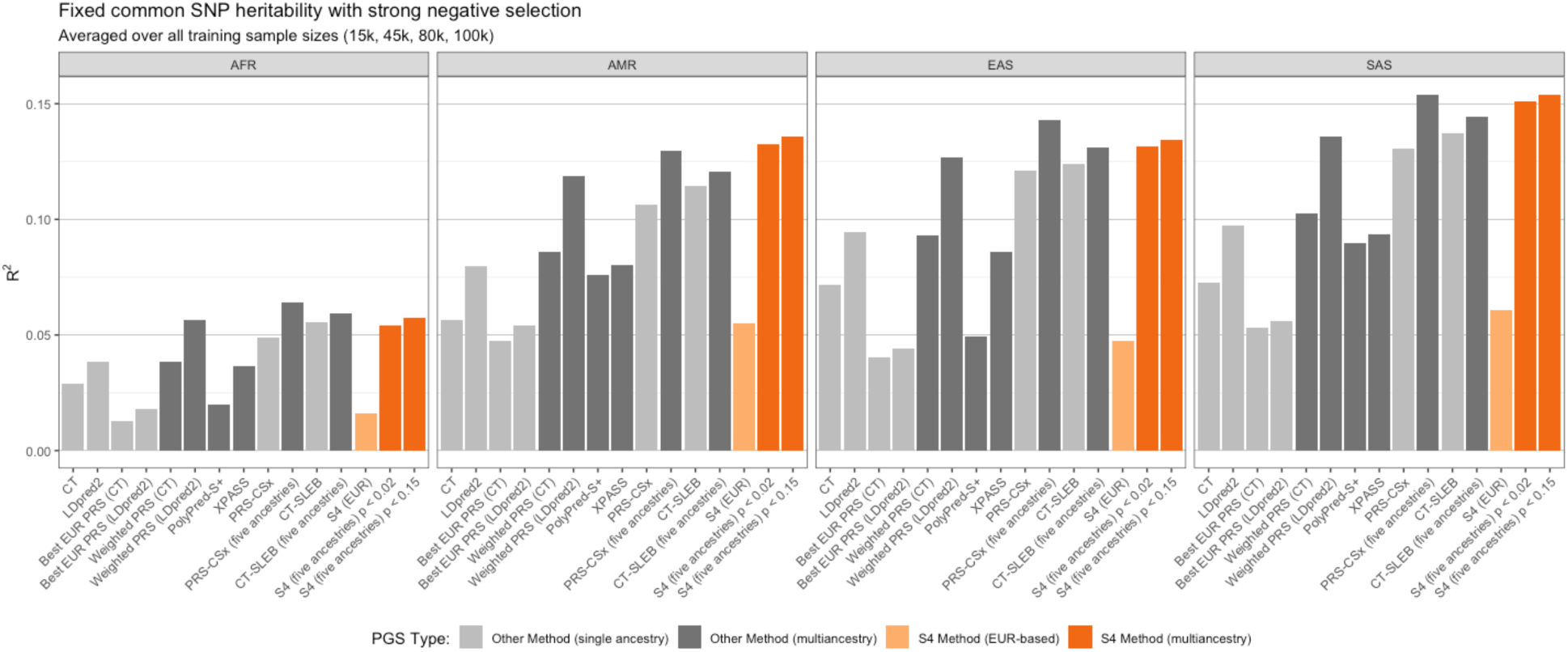
simulation results, averaged over all training sample sizes

While all PRS constructions showed lower performance in AFR target ancestries (*Figs. 2-3*), multi-ancestry methods achieved a substantial improvement over single-ancestry and EUR-based constructions in AFR prediction. Moreover, this gap in accuracy decreased as the training sample size grew. At a training sample size of 15,000, prediction *R*^2^was 54% lower in AFR target sample than the next lowest accuracy ancestry (AFR: 0.036 vs. EAS: 0.078). While this difference dropped to a 36% decrease at a training sample size of 100,000 (AMR: 0.136 vs. AFR: 0.087), a substantial performance gap remained even at the largest tested sample size.

**Figure 3:**
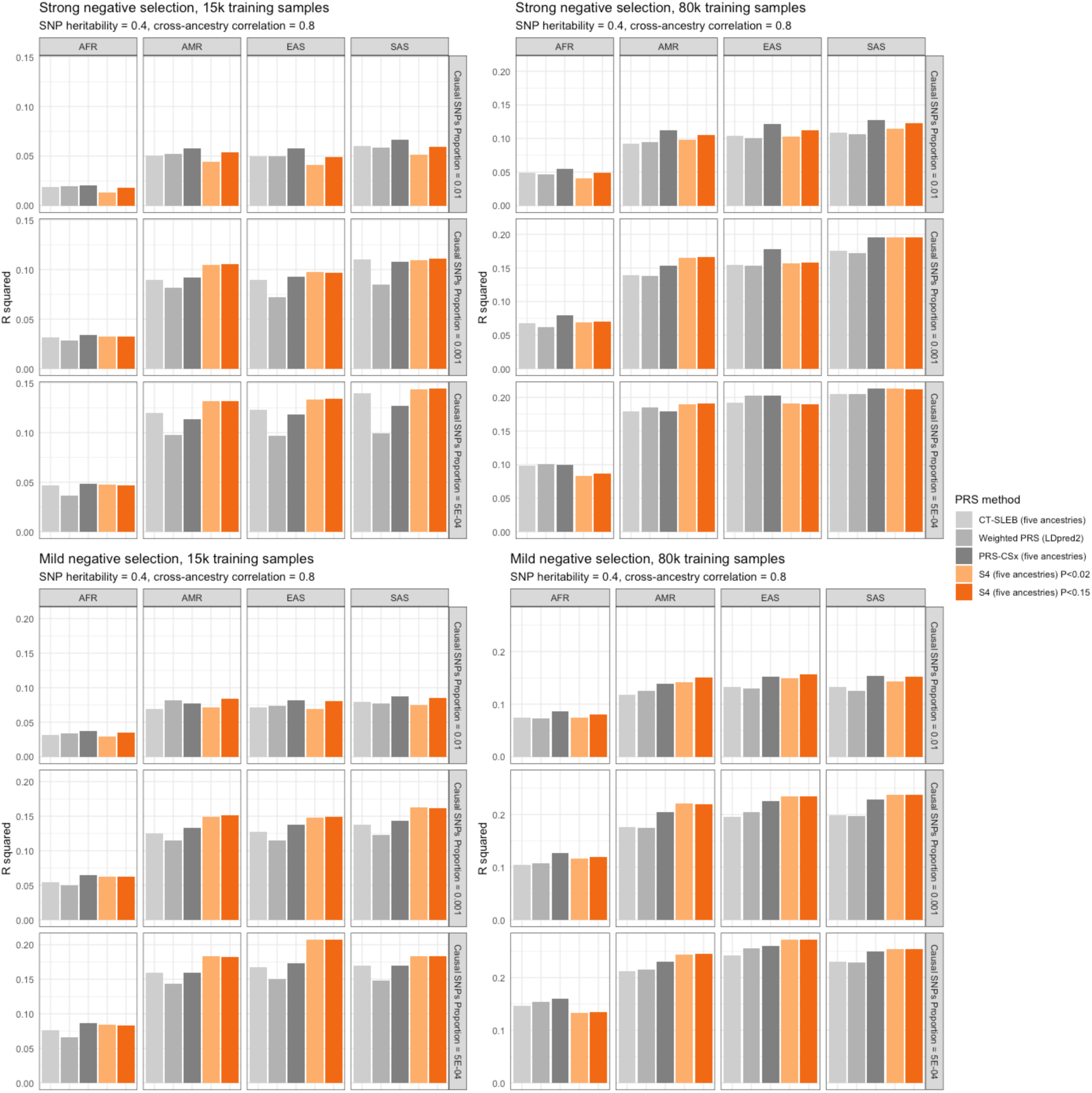
Multi-ancestry methods, stratified by genetic architecture sparsity and ancestry

Both tested *p*-value cutoffs (0.02 and 0.15) for S4-Multi performed comparably overall, with a mean accuracy increase of just 2.5% for the larger model averaged over all simulation runs (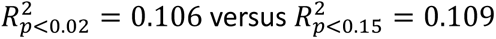). This suggests that the specific threshold is generally not a make-or-break decision in implementing S4-Multi in most cases. The performance gain from the higher cutoff was greatest when applied to a highly polygenic setting (*Fig. S4*), achieving a 10% increase in accuracy averaged over all such runs. In all other polygenicity settings, the performance difference was nearly negligible (< 1%). The maximum difference occurred under fixed common SNP heritability with mild negative selection and a training sample size of 15,000 and causal SNP proportion of 0.01, with the higher threshold model achieving a 16% increase in accuracy (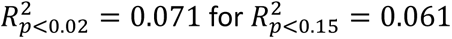).

This was the most challenging prediction setting for S4-Multi in general—low degree of polygenicity and small sample size—so it is somewhat intuitive that this is where the additional SNPs in the larger model provided the most benefit. However, this modest overall change in accuracy over the genetic architectures and sample sizes tested gives some indication that S4-Multi retains relatively strong prediction accuracy as the number of available SNPs decreases.

*Table 1* records the number of simulation runs in which each method was the top performer over the 60 total runs in each target ancestry. These span all tested proportions of causal SNPs, genetic architectures, and sample sizes. S4-Multi and PRS-CSx consistently outperformed all other tested methods overall, with S4-Multi tending to perform comparably in AMR, EAS, and SAS ancestries, and PRS-CSx ranking consistently higher in AFR prediction. S4-Multi improved upon LDpred2 and CT-SLEB across EAS, SAS and EUR validation data irrespective of underlying genetic architecture and training sample size. While PRS-CSx achieved top performance in 122/240 = 51% of runs overall versus 99/240 = 41% for S4-Multi, this difference was primarily driven by prediction in AFR ancestries.

**Table 1:** The number of simulation runs (out of all 60 different sample constructions per ancestry) in which each PRS was the top performer is shown. All other tested models had zero runs in which they were the top performer.

| PRS Construction: | Target ancestry: |  |  |  | Total: |
| --- | --- | --- | --- | --- | --- |
|  | AFR: | AMR: | EAS: | SAS: |  |
| CT-SLEB | 2 | 1 | 0 | 1 | 4 |
| LDpred2 | 1 | 0 | 0 | 0 | 1 |
| LDpred2 (weighted) | 5 | 2 | 1 | 1 | 9 |
| PRS-CSx | 48 | 21 | 30 | 23 | 122 |
| S4-Multi | 2 | 36 | 28 | 33 | 99 |
| XPASS | 2 | 0 | 1 | 2 | 5 |
| **Total:** | 60 | 60 | 60 | 60 | 240 |

*Fig. 3* highlights that the underlying genetic architecture had little impact on the relative accuracy of PGS conctructions at the smallest tested training sample size (*n* = 15,000). At larger training sample sizes (*n* = 80,000), S4-Multi showed modest improvement under mild negative selection pressure among AMR and EAS validation sets relative to other multi-ancestry methods (PRS-CSx, LDpred2, CT-SLEB). While there were marginal changes as causal SNP proportion and sample size varied, there was generally limited difference between methods. While the S4-Multi methods showed some improvement under mild negative selection pressure, genetic architecure had limited impact on relative accuracy of PRS constructions in general, with all methods performing slightly better under strong negative selection for a given sample size. The strongest overall trends were the superior performance of multi-ancestry methods over single-ancestry and EUR-only counterparts, and remaining performance gap for prediciton in AFR ancestries, particularly at lower traninng sample sizes.

### Biobank data testing

We evaluated S4-Multi, LDpred2, and PRS-CSx on available biobank data. All models were trained, tuned, and validated on available data from UK Biobank (UKB), FinnGen, Biobank Japan (BBJ), *All of Us* (AoU), and the Global Biobank Initiative (GBB) to predict five complex traits: type 2 diabetes, breast cancer, colorectal cancer, asthma, and stroke (either ischemic or hemorrhagic). Sample sizes for each phenotype and cohort are available in *Table S4*. During PRS-CSx model development, separate models were fit on each available training ancestry before regressing the results on the target ancestry to produce the final PGM for testing.Individuals were grouped based on genetically inferred ancestry computed from principal component (PC) analysis and classification algorithm within each biobank.

Implementing the S4-Multi PGM requires three independent datasets for model development, tuning, and validation. Model development was performed on two settings: (1) using a consortium of large-scale GWAS summary statistics from UK Biobank, FinnGen, and Biobank Japan, spanning all available ancestries (AFR, EAS, EUR, and SAS), and (2) using EUR data only. Due to the limited sample size for colorectal cancer phenotype, model development data were available only for EAS and EUR ancestries. Model tuning was performed on available EUR genotype data, with UKB and FinnGen runs separated to control for differences in cohort population structure. Model validation was conducted on AFR, AMR, EAS, and EUR ancestries using data from available biobanks (see *Table 3*). Case and control counts for the validation sets of each phenotype and biobank source are available in the supplementary materials (*Table S3*). Asthma and stroke development and testing was performed on a consortium of biobank data from the Global Biobank Initiative^39^.

Full results for all runs on biobank data are presented in *Table 2*. Model performance was measured with the log odds ratio per standard deviation of PGS (log(OR) per SD). *χ*^2^statistics from the corresponding likelihood ratio tests as well as results for models trainined only on EUR ancestry data are provided in the summplementary materials (*Tables S2-S3*). In subsequent discussion, we consider log(OR) per SD as the performance metric. Multi-ancestry methods trained on all available development ancestries showed the strongest performance in type 2 diabetes prediction across all target ancestries and PGM methods (mean = 0.570), with all results falling in the interval [0.254, 0.948]. Prediction in breast cancer (mean = 0.452, interval = [0.269, 0.623]), asthma (mean = 0.369, interval = [0.146, 0.530]), colorectal cancer (mean = 0.344, interval = [0.136, 0.551]), and stroke (mean = 0.090, interval = [-0.175, 0.204]) was more challenging with a substantial dropoff in overall performance for the latter.

**Table 2:** Full-ancestry data used in model development consist of a consortium of available data from UK Biobank (UKB), FinnGen, and Biobank Japan (BBJ). The top performing PGM is highlighted for each combination of development and test set. Models for asthma and stroke were developed on full-ancestry data from Global Biobank (GBB). Results for models developed only on EUR data are available in the supplementary materials (Table S2).

| Type 2 Diabetes |  |  |  |  |  |  |
| --- | --- | --- | --- | --- | --- | --- |
| MODEL DEVELOPMENT |  | MODEL TESTING |  |  |  |  |
| Development set | Tuning set (EUR) | Validation set |  | Performance (log(OR) per SD) |  |  |
|  |  | Ancestry | Cohort | S4 | LDpred2 | PRS-CSx |
| EAS, EUR, SAS | FinnGen | AFR | AoU | 0.256 | 0.254 |  |
|  | UKB |  |  | 0.257 | 0.270 |  |
|  | FinnGen |  | UKB | 0.343 | 0.347 | 0.399 |
|  | FinnGen | AMR | AoU | 0.512 | 0.379 |  |
|  | UKB |  |  | 0.521 | 0.413 |  |
|  | FinnGen | EAS | AoU | 0.533 | 0.588 | 0.594 |
|  | UKB |  |  | 0.542 | 0.590 | 0.603 |
|  | FinnGen |  | BBJ | 0.948 | 0.813 | 0.936 |
|  | UKB |  |  | 0.919 | 0.804 | 0.904 |
|  | FinnGen |  | UKB | 0.587 | 0.553 | 0.491 |
|  | FinnGen | EUR | AoU | 0.537 | 0.568 | 0.566 |
|  | UKB |  |  | 0.532 | 0.569 | 0.571 |
|  | FinnGen |  | UKB | 0.717 | 0.673 | 0.688 |
|  | UKB |  | FinnGen | 0.661 | 0.604 | 0.617 |

| Breast Cancer |  |  |  |  |  |  |
| --- | --- | --- | --- | --- | --- | --- |
| MODEL DEVELOPMENT |  | MODEL TESTING |  |  |  |  |
| Development set | Tuning set (EUR) | Validation set |  | Performance (log(OR) per SD) |  |  |
|  |  | Ancestry | Cohort | S4 | LDpred2 | PRS-CSx |
| AFR, EAS, EUR | FinnGen | AFR | AoU | 0.344 | 0.354 |  |
|  | UKB |  |  | 0.351 | 0.379 |  |
|  | FinnGen | AMR | AoU | 0.388 | 0.404 |  |
|  | UKB |  |  | 0.391 | 0.406 |  |
|  | FinnGen | EAS | AoU | 0.417 | 0.347 | 0.269 |
|  | UKB |  |  | 0.413 | 0.363 | 0.269 |
|  | FinnGen |  | BBJ | 0.457 | 0.456 | 0.432 |
|  | UKB |  |  | 0.454 | 0.460 | 0.432 |
|  | FinnGen | EUR | AoU | 0.532 | 0.550 | 0.523 |
|  | UKB |  |  | 0.530 | 0.550 | 0.523 |
|  | FinnGen |  | UKB | 0.565 | 0.552 | 0.533 |
|  | UKB |  | FinnGen | 0.623 | 0.619 | 0.577 |

| Colorectal Cancer |  |  |  |  |  |  |
| --- | --- | --- | --- | --- | --- | --- |
| MODEL DEVELOPMENT |  | MODEL TESTING |  |  |  |  |
| Development set | Tuning set (EUR) | Validation set |  | Performance (log(OR) per SD) |  |  |
|  |  | Ancestry | Cohort | S4 | LDpred2 | PRS-CSx |
| EAS, EUR | FinnGen | AFR | AoU | 0.136 | 0.156 |  |
|  | UKB |  |  | 0.149 | 0.156 |  |
|  | FinnGen | AMR | AoU | 0.195 | 0.219 |  |
|  | UKB |  |  | 0.206 | 0.219 |  |
|  | FinnGen | EAS | AoU | 0.382 | 0.387 |  |
|  | UKB |  |  | 0.368 | 0.387 |  |
|  | FinnGen | EUR | AoU | 0.386 | 0.376 | 0.344 |
|  | UKB |  |  | 0.380 | 0.376 | 0.344 |
|  | FinnGen |  | UKB | 0.512 | 0.504 | 0.475 |
|  | UKB |  | FinnGen | 0.551 | 0.547 | 0.513 |

| Asthma |  |  |  |  |  |  |
| --- | --- | --- | --- | --- | --- | --- |
| MODEL DEVELOPMENT |  | MODEL TESTING |  |  |  |  |
| Development set | Tuning set (EUR) | Validation set |  | Performance (log(OR) per SD) |  |  |
|  |  | Ancestry | Cohort | S4 | LDpred2 | PRS-CSx |
| AFR, AMR, EAS, EUR, SAS | GBB | AFR | AoU | 0.252 | 0.236 | 0.197 |
|  |  |  | UKB | 0.260 | 0.213 |  |
|  |  | AMR | AoU | 0.387 | 0.320 | 0.146 |
|  |  | EAS | AoU | 0.527 | 0.530 | 0.485 |
|  |  |  | UKB | 0.237 | 0.222 |  |
|  |  | EUR | AoU | 0.373 | 0.390 | 0.365 |
|  |  |  | UKB | 0.505 | 0.500 | 0.480 |

| Stroke (ischemic and hemorrhagic) |  |  |  |  |  |  |
| --- | --- | --- | --- | --- | --- | --- |
| MODEL DEVELOPMENT |  | MODEL TESTING |  |  |  |  |
| Development set | Tuning set (EUR) | Validation set |  | Performance (log(OR) per SD) |  |  |
|  |  | Ancestry | Cohort | S4 | LDpred2 | PRS-CSx |
| AMR, AFR, EAS, EUR | GBB | AFR | AoU | 0.088 | 0.073 | 0.015 |
|  |  |  | UKB | 0.180 | 0.190 |  |
|  |  | AMR | AoU | -0.112 | -0.105 | -0.175 |
|  |  | EAS | AoU | 0.046 | 0.155 | -0.094 |
|  |  |  | UKB | 0.104 | 0.118 |  |
|  |  | EUR | AoU | 0.139 | 0.138 | 0.107 |
|  |  |  | UKB | 0.198 | 0.191 | 0.171 |

Mirroring the simulation results, all methods tested showed improved performance when trained on all available ancestries versus only utilizing EUR training data (*Table* S2). Multi-ancestry PGMs improved performance by 13% on average (in log(OR) / SD) relative to their single-ancestry counterparts across all methods, phenotypes, and ancestries. The overall performance gains for the multi-ancestry methods were greatest for type 2 diabetes and breast cancer prediction (0.085 and 0.032, respectively), with more modest increases in colorectal cancer (0.024). EUR-only models were not trained for asthma and stroke. Overall performance gains were largest in EAS (28% greater) and AFR (10% greater) prediction, though there were still modest improvements in AMR (5.6% greater) and EUR (3.4% greater) origin populations. Across all three phenotypes, we again found substantially reduced accuracy in AFR ancestries. In both breast cancer and type 2 diabetes prediction, the worst-performing EUR-based PGM in non-AFR ancestries outperformed the best-performing multi-ancestry PGM in AFR ancestries. This highlights the lingering challenges of PGS prediction in AFR-origin populations despite these improvements.

The best-performing PGM method varied considerably both with target ancestry and phenotype (*Fig. 4*). For example, while PRS-CSx was the strongest predictor of type 2 diabetes in EAS individuals, it was the least accurate of the three tested models for predicting breast cancer in the EAS target population. However, with some exceptions such as S4’s strong performance predicting breast cancer in EAS populations, the three multi-ancestry methods tended to achieve similar results in cases where all models were tested. We found similar results in cases when only S4 and LDpred2 were tested, apart from S4’s strong performance predicting type 2 diabetes in AMR-origin populations. Within each phenotype-ancestry-cohort combination, the worst-performing PGM was only 13% less accurate (in log(OR) per SD) than the top-performing model averaged over type 2 diabetes, breast cancer, colorectal cancer, and asthma tests. Performance in stroke prediction was more variable with a mean 53% decrease between the top-performing and worst-performing PGM. However, all methods suffered from considerably lower accuracy for stroke relative to other phenotypes. Among the non-stroke diseases tested, the maximum difference occurred in asthma prediction in AMR-origin individuals from the *AoU* cohort, where PRS-CSx was 62% less accurate than the top-preforming model (S4).

**Figure 4:**
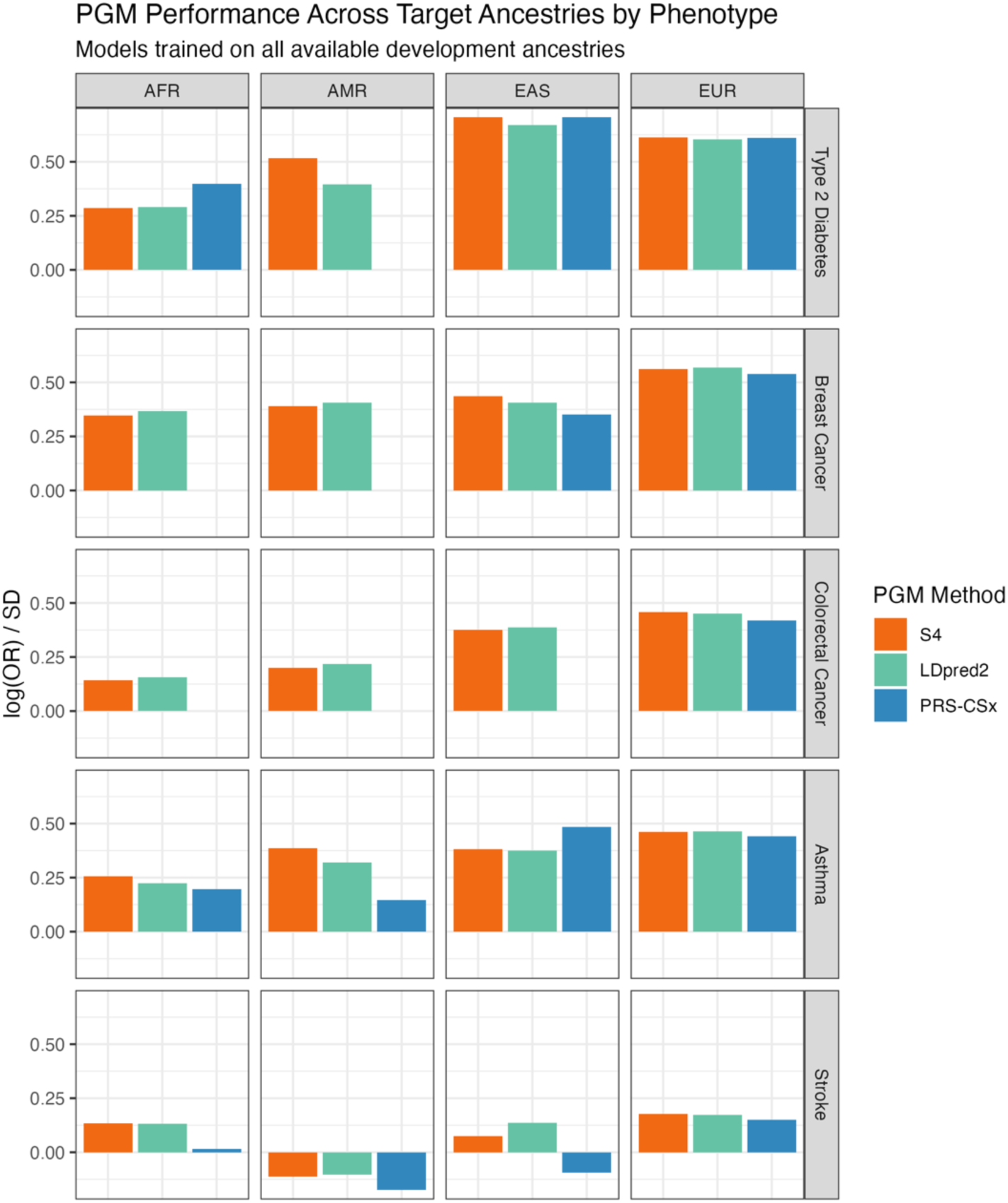
The best-performing PGM method varied substantially with both phenotype and ancestry. Model performance was averaged over all biobank runs for a given phenotype-ancestry combination. Note that PRS-CSx was not fit on all phenotype-ancestry combinations.

All tested models showed diminished accuracy predicting stroke relative to other phenotypes across all tested ancestries. Within each tested group of ancestries, stroke prediction was on average 84% less accurate than the next lowest phenotype. Notably, all three methods produced negative log(OR) per SD results in the tested AMR-origin population. This implies that both models produced lower scores on average in controls than in cases. We give these results particular consideration here.

Using genetic principal components (PCs) computed from a set of LD-pruned (*r*^2^ < 0.1) autosomal SNPs (provided by *All of Us*), we examined the correlation between the PGSs and the first four PCs. We find that the tested PGSs in AMR-origin individuals are weakly correlated with each of the first four PCs (*Table S5, Fig. S5*), and that cases and controls are distributed differently along those PCs. This suggest that differences in the population structure between cases and controls may confound the association between PGS and phenotype. Adjusting for the first 16 PCs in the logistic regression model returns positive log(OR) per SD estimates for tested PGSs, suggesting that population structure is a likely confounder here. Similar investigation into type 2 diabetes results also reveals correlations and distributional differences between AMR-origin cases and controls along genetic PCs, albeit to a lesser degree (*Fig. S6*). However, since all PGM methods were more accurate in general at predicting type 2 diabetes, the tested models nonetheless produce a positive result (in log(OR) per SD) for AMR prediction. We present the results from the unadjusted models in *Table 2* and *Fig. 4* to maintain consistency across simulation test, biobank tests, and replicated analyses, though the potential for population structure confounding in highly admixed populations should be noted and likely warrants specific consideration for future work.

## Discussion

In this paper, we presented S4-Multi, an extension of the S4 PGM that leverages data across multiple ancestries for improved cross-ancestry prediction. We compared its performance with a range of single- and multi-ancestry PGM methods for prediction across the ancestry continuum in both simulated and biobank data. We find that S4-Multi makes significant performance gains in cross-ancestry prediction compared to its single-ancestry implementation. Moreover, S4-Multi achieves state-of-the-art performance both in simulated data and large-scale biobank data across type 2 diabetes, breast cancer, and colorectal cancer prediction tasks. Simulation results indicate that S4’s retains strong performance at smaller sample sizes relative to alternative multi-ancestry PGS constructions. That S4-Multi achieved top or near-top performance in both the simulation benchmarking and real-world biobank prediction tests is encouraging for its robustness across a range of settings, phenotypes, and sample characteristics.

We found limited differences in performance between the top multi-ancestry PGS methods in general, with the best-performing model being highly dependent on phenotype, target ancestry, and genetic architecture. This aligns with previous findings that no PGS method is uniformly superior in all contexts^34^. Our biobank tests lend credence to this in real world settings, with relative performance between PGS methods varying considerably between phenotypes. This suggests that disease-specific and ancestry-specific genetic architecture plays a role in differentiating PGS performance, matching prior work^40^. Evaluating multiple PGM constructions is likely prudent for clinical use to identify the best-performing model for specific prediction tasks and target populations.

We see evidence that differences in underlying population structure within classified ancestries may confound the underlying association between the PGS and phenotype, as indicated by the discussion of AMR stroke prediction above. This is consistent with prior work finding PGSs for a range of traits to correlate with geographic population distribution, even in cases with relatively homogenous populations^41^. This trend being most pronounced in AMR target populations suggests that prediction in highly admixed populations likely warrants particular attention moving forward, as well as supporting the need to consider PGS performance along the continuum of genetic ancestries rather than simply in discrete ancestry groups^19,42^.

A central limitation in model development and testing remains the smaller available sample sizes from AFR, AMR, and SAS origin populations in GWAS data. While projects aiming to diversify participation such as *All of Us* enabled model testing on observed data across the ancestry continuum, training and validation sample sizes remain limited for non-EUR origin populations. This was particularly challenging for lower-prevalence diseases like colorectal cancer, where sufficient training data was only available for training and development on EUR- and EAS-origin populations in our tests. Continuing to diversify GWAS participation is critical for multi-ancestry PGM development and evaluation moving forward. Given the current magnitude of the EUR overrepresentation in GWAS and limited available training data, strong performance at lower training sample sizes is likely to be a meaningful differentiator of PGMs for practical use. This is especially true for low-prevalence disease prediction, which exacerbates sample size issues for underrepresented populations in GWAS.

Both simulation and biobank results suggest that while multi-ancestry methods make meaningful inroads for risk stratification in AFR target ancestries, this remains a challenging setting for PGS prediction. While increasing training sample size in simulation tests reduced this accuracy drop in AFR target ancestries to a degree, a meaningful performance gap remained at the largest tested training sample size (*n* = 100,000). This suggests that while continuing to diversify GWAS participation will be able to attenuate this somewhat, further model refinement for multi-ancestry methods is likely necessary to match prediction accuracy in EUR-origin populations.

A further benefit of S4 was its strong performance despite including fewer variants in the final model across all five tested phenotypes in biobank tests (*Fig. 5, Table S6*). Relative to the next sparsest PGM, S4 used from 9% fewer (asthma) to 77% fewer (colorectal cancer) variants. This is an encouraging indication that S4 retains good prediction accuracy with fewer available SNPs, though this warrants further investigation verifying the robustness of PGM methods in clinical settings where available genomic data are in the hundreds or thousands of variants are available rather than the millions considered here.

**Figure 5:**
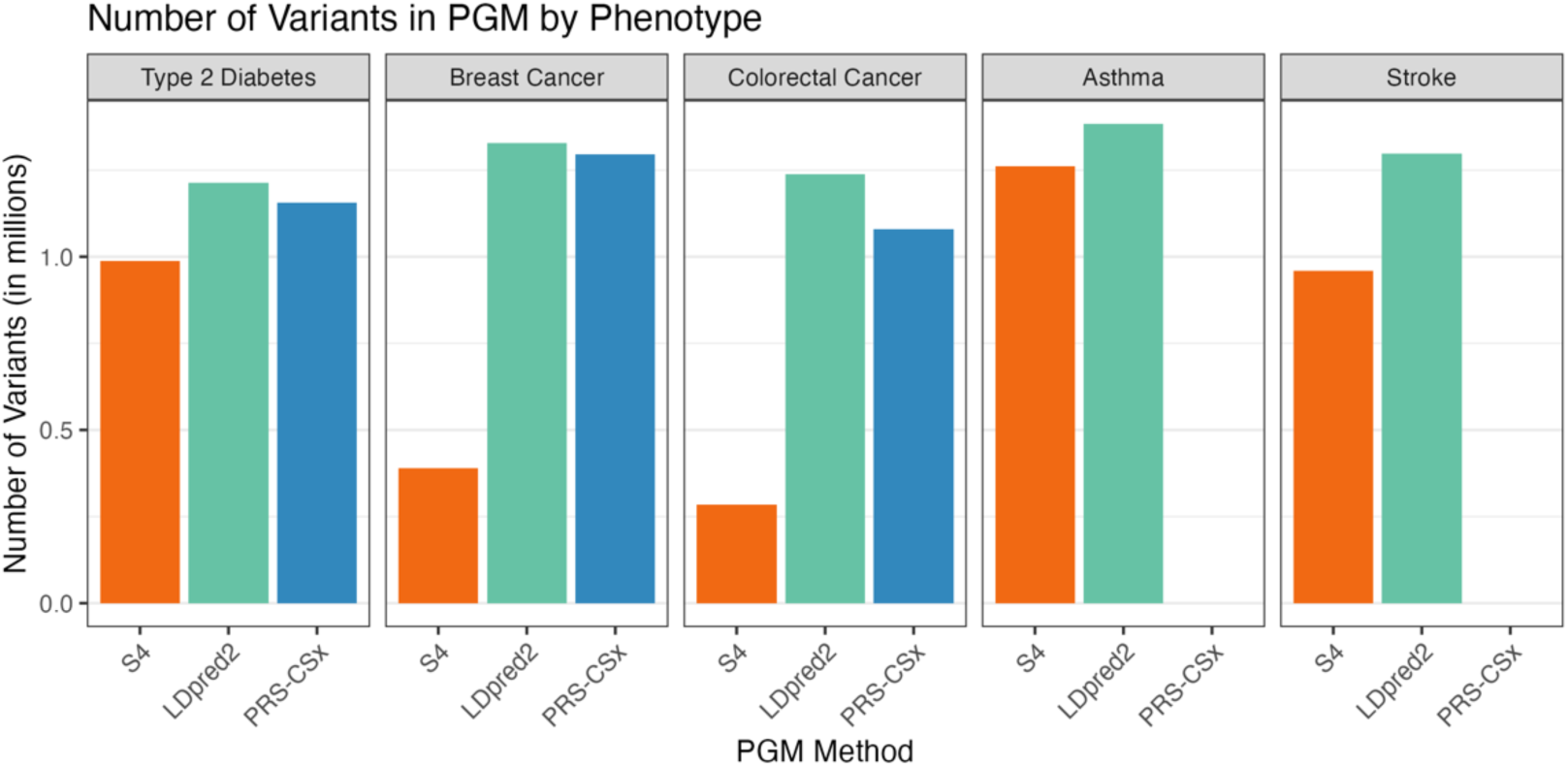
S4 achieves state-of-the-art performance with fewer variants in final PGM, suggesting potential for robust performance in settings with fewer available variants

In conclusion, we have presented a new multi-ancestry PGS construction generated from GWAS and genotype data from diverse populations. S4-Multi achieves strong performance across a range of phenotypes for improved cross-ancestry prediction.

## Data Availability

All data used in this analysis is publicly available online. The simulated data is identical to that used in Zhang et al 2023 and is publicly available on GitHub (https://github.com/andrewhaoyu/multi_ethnic). Biobank tests utilized both summary statistics for model development and individual-level genomic data for testing. Individual-level genomic data used in this analysis is publicly accessible after registration with the following biobanks: All of Us (v7 release) (https://www.researchallofus.org/), UK Biobank (https://www.ukbiobank.ac.uk/), FinnGen (https://www.finngen.fi/en), Biobank Japan (https://biobankjp.org/en/), and the Global Biobank Meta-analysis Initiative (https://www.globalbiobankmeta.org/). GWAS summary statistics by phenotype are available here: breast cancer (http://bcac.ccge.medschl.cam.ac.uk), colorectal cancer (https://www.ebi.ac.uk/gwas/studies/GCST90255677,
https://www.ebi.ac.uk/gwas/publications/36539618,
https://www.ebi.ac.uk/gwas/studies/GCST90129505), type 2 diabetes (https://diagram-consortium.org/downloads.html, https://t2d.hugeamp.org), asthma (https://gbmi-sumstats.s3.amazonaws.com/Asthma_Bothsex_afr_inv_var_meta_GBMI_052021_nbbkgt1.txt.gz, https://gbmi-sumstats.s3.amazonaws.com/Asthma_Bothsex_amr_inv_var_meta_GBMI_052021_nbbkgt1.txt.gz, https://gbmi-sumstats.s3.amazonaws.com/Asthma_Bothsex_eas_inv_var_meta_GBMI_052021_nbbkgt1.txt.gz, https://gbmi-sumstats.s3.amazonaws.com/Asthma_Bothsex_eur_inv_var_meta_GBMI_052021_nbbkgt1.txt.gz, https://gbmi-sumstats.s3.amazonaws.com/Asthma_Bothsex_sas_inv_var_meta_GBMI_052021_nbbkgt1.txt.gz, https://gbmi-sumstats.s3.amazonaws.com/Asthma_Bothsex_leave_UKBB_inv_var_meta_GBMI_052021.txt.gz), stroke (https://gbmi-sumstats.s3.amazonaws.com/Stroke_Bothsex_afr_inv_var_meta_GBMI_052021_nbbkgt1.txt.gz, https://gbmi-sumstats.s3.amazonaws.com/Stroke_Bothsex_amr_inv_var_meta_GBMI_052021_nbbkgt1.txt.gz, https://gbmi-sumstats.s3.amazonaws.com/Stroke_Bothsex_amr_inv_var_meta_GBMI_052021_nbbkgt1.txt.gz, https://gbmi-sumstats.s3.amazonaws.com/Stroke_Bothsex_eur_inv_var_meta_GBMI_052021_nbbkgt1.txt.gz, https://gbmi-sumstats.s3.amazonaws.com/Stroke_Bothsex_leave_UKBB_inv_var_meta_GBMI_052021.txt.gz)

https://www.researchallofus.org/

https://www.ukbiobank.ac.uk/

https://www.finngen.fi/en

https://biobankjp.org/en/

https://www.globalbiobankmeta.org/

http://bcac.ccge.medschl.cam.ac.uk

https://www.ebi.ac.uk/gwas/studies/GCST90255677

https://www.ebi.ac.uk/gwas/publications/36539618

https://www.ebi.ac.uk/gwas/studies/GCST90129505

## Acknowledgements

We thank *All of Us* participants for their contributions. We also thank the *All of Us* Research Program for making available the participant data examined in this study.

## Data availability

The simulated data used in this analysis is identical to that used in Zhang et al 2023^34^ and is publicly available on GitHub. Biobank tests utilized both summary statistics for model development and individual-level genomic data for testing. Sources for summary statistics are provided in the table below. Individual-level genomic data used in this analysis is publicly accessible after registration with the following biobanks: *All of Us* (v7 release) (https://www.researchallofus.org/), UK Biobank (https://www.ukbiobank.ac.uk/), FinnGen (https://www.finngen.fi/en), Biobank Japan (https://biobankjp.org/en/), and the Global Biobank Meta-analysis Initiative (https://www.globalbiobankmeta.org/).

| Phenotype: | GWAS summary statistic sources: |
| --- | --- |
| Breast cancer | EUR summary statistics from Michailidou et al 2017<sup>43</sup> |
| Colorectal cancer | GWAS catalog numbers: GCST90255677, GCST90255676, GCST90129505 |
| Type 2 diabetes | Available from DIAGRAM Consortium (https://diagram-consortium.org/downloads.html)<br>GWAS meta-analysis summary statistics from Mahajan et al 2022<sup>44</sup> |
| Asthma | Download links:<br>https://gbmi-sumstats.s3.amazonaws.com/Asthma_Bothsex_afr_inv_var_meta_GBMI_052021_nbbkgt1.txt.gz<br>https://gbmi-sumstats.s3.amazonaws.com/Asthma_Bothsex_amr_inv_var_meta_GBMI_052021_nbbkgt1.txt.gz<br>https://gbmi-sumstats.s3.amazonaws.com/Asthma_Bothsex_eas_inv_var_meta_GBMI_052021_nbbkgt1.txt.gz |
|  | • https://gbmi-sumstats.s3.amazonaws.com/Asthma_Bothsex_eur_inv_var_meta_GBMI_052021_nbbkgt1.txt.gz<br>• https://gbmi-sumstats.s3.amazonaws.com/Asthma_Bothsex_sas_inv_var_meta_GBMI_052021_nbbkgt1.txt.gz<br>• https://gbmi-sumstats.s3.amazonaws.com/Asthma_Bothsex_leave_UKBB_inv_var_meta_GBMI_052021.txt.gz |
| Stroke | Download links:<br>• https://gbmi-sumstats.s3.amazonaws.com/Stroke_Bothsex_afr_inv_var_meta_GBMI_052021_nbbkgt1.txt.gz<br>• https://gbmi-sumstats.s3.amazonaws.com/Stroke_Bothsex_amr_inv_var_meta_GBMI_052021_nbbkgt1.txt.gz<br>• https://gbmi-sumstats.s3.amazonaws.com/Stroke_Bothsex_eas_inv_var_meta_GBMI_052021_nbbkgt1.txt.gz<br>• https://gbmi-sumstats.s3.amazonaws.com/Stroke_Bothsex_eur_inv_var_meta_GBMI_052021_nbbkgt1.txt.gz<br>• https://gbmi-sumstats.s3.amazonaws.com/Stroke_Bothsex_leave_UKBB_inv_var_meta_GBMI_052021.txt.gz |

## Supplementary Materials

**Figure S1:**
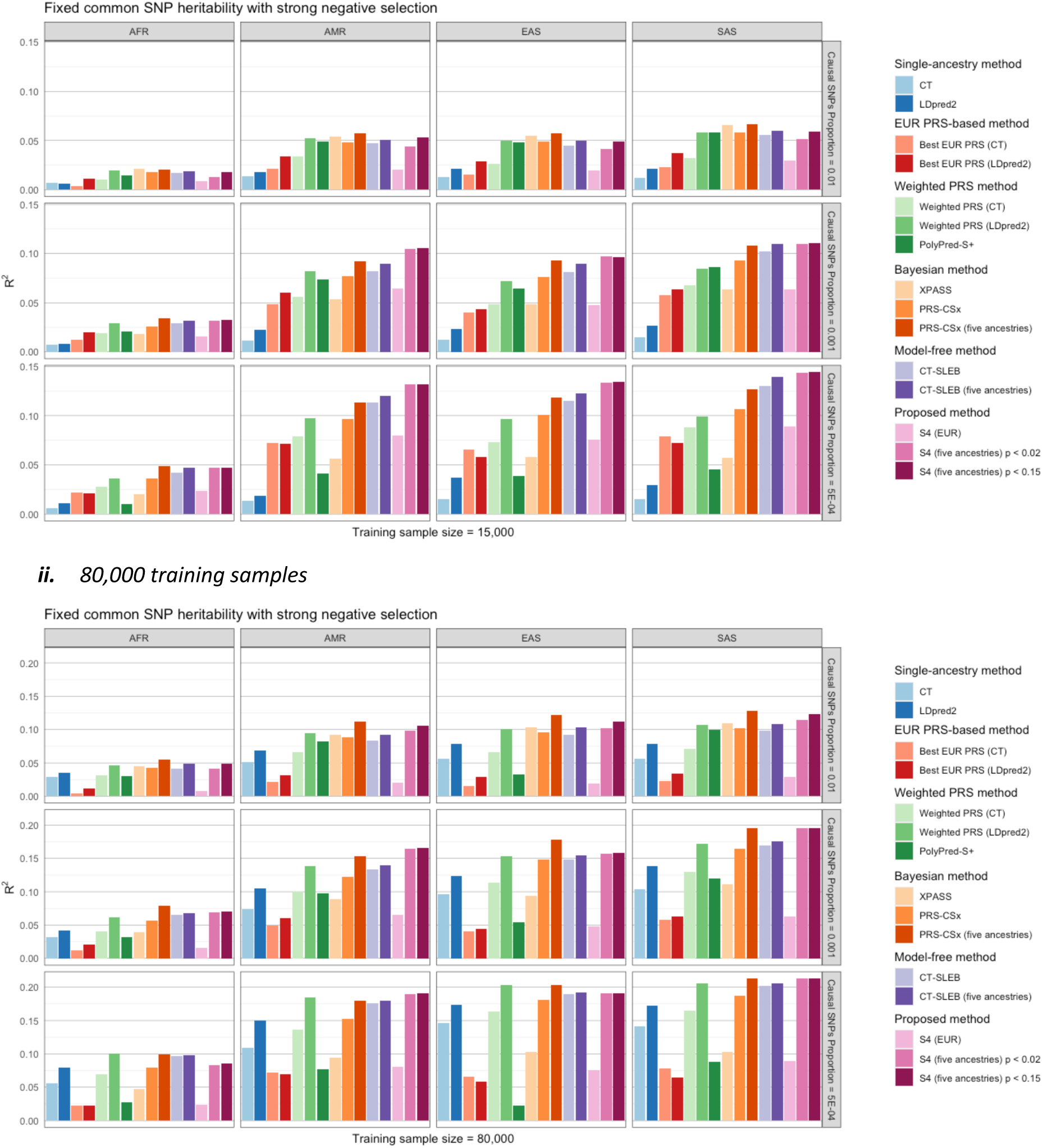
simulation results, stratified by target ancestries and causal SNP proportion

**Figure S2:**
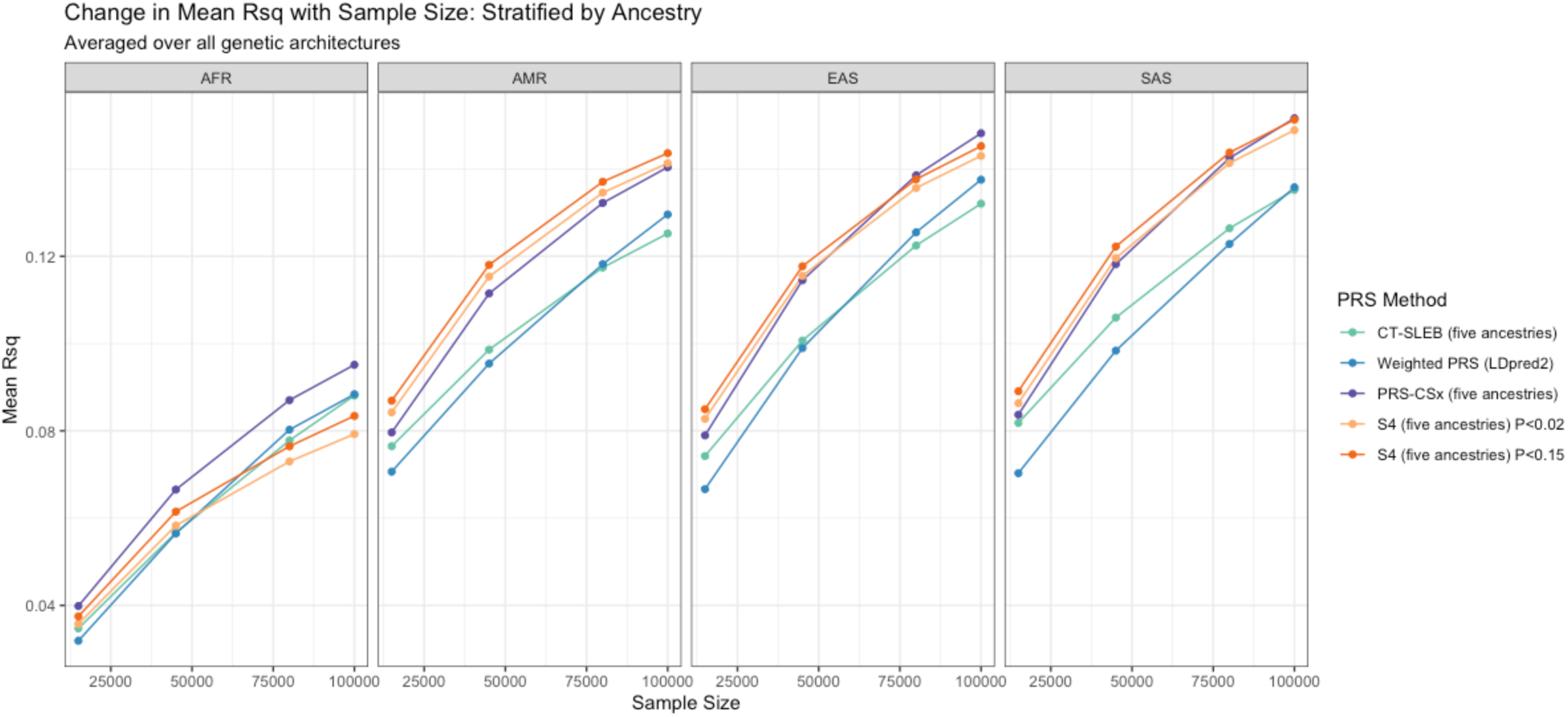
Simulated data prediction accuracy stratified by ancestries and averaged over all runs at each sample size-ancestry combination

**Figure S3:**
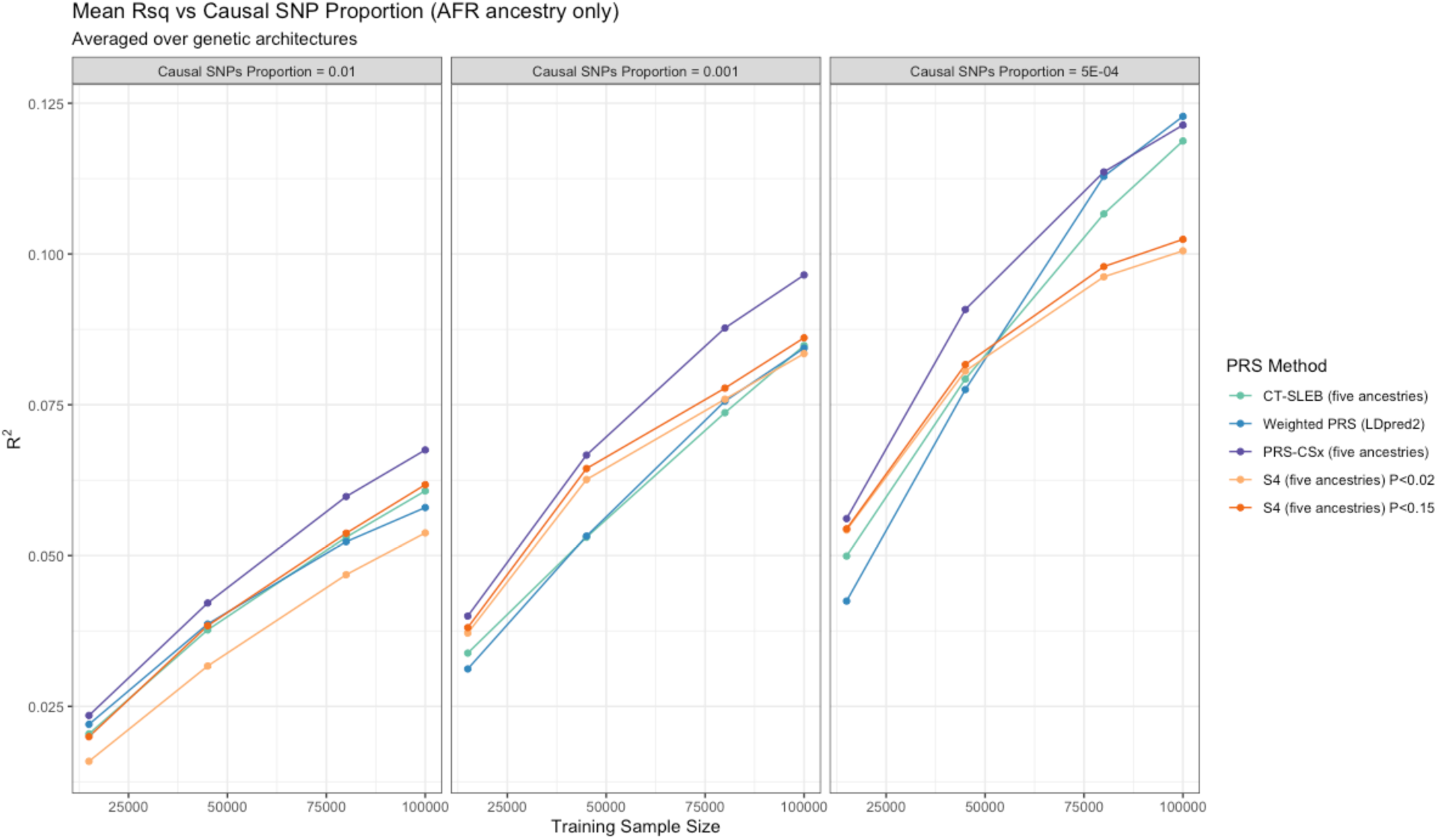
Simulation results on only AFR-ancestries prediction, stratified by causal SNP proportion

**Figure S4:**
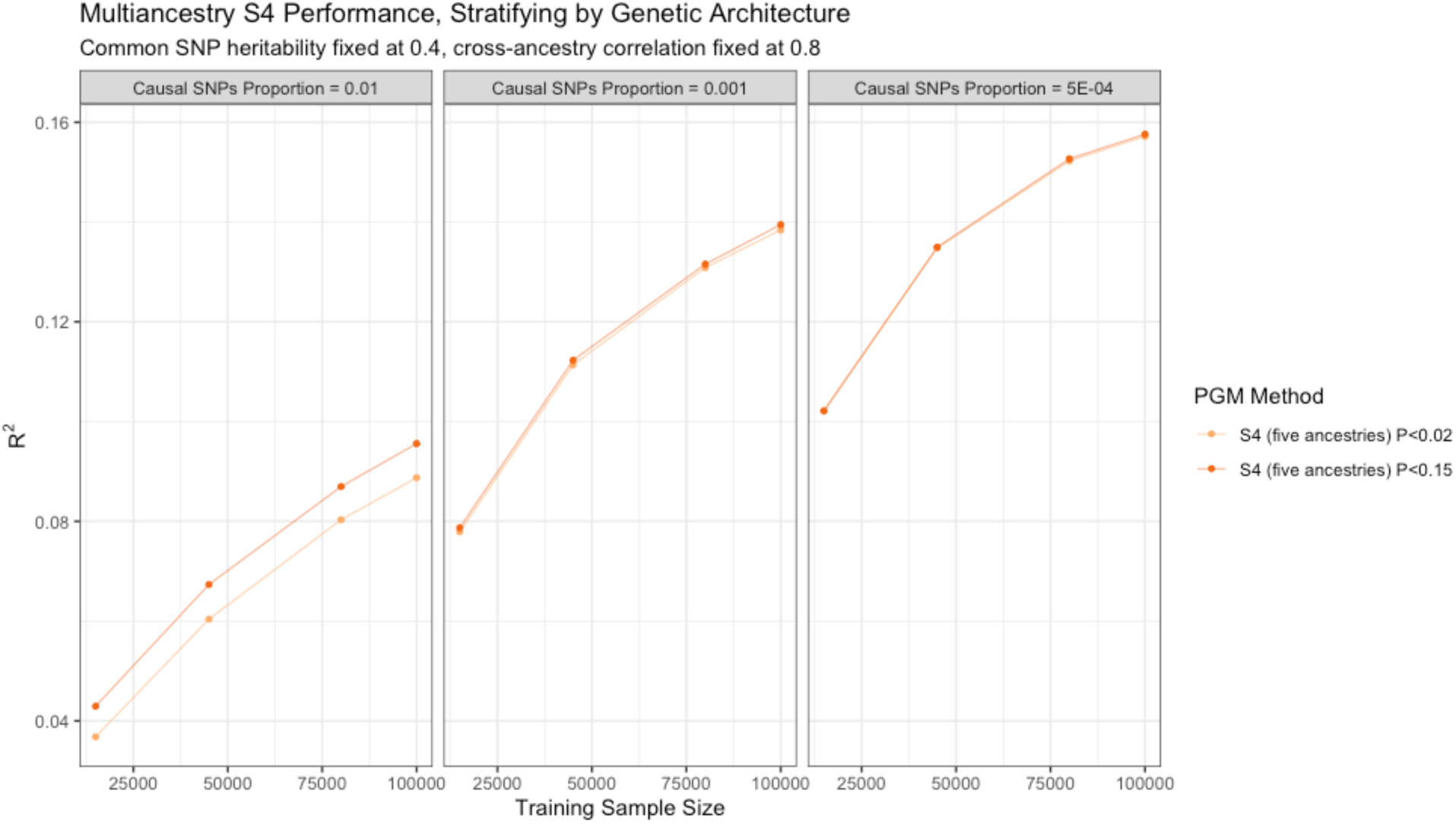
Comparing performance of different p-value cutoffs for S4-Multi across genetic architectures and sample sizes

**Figure S5:**
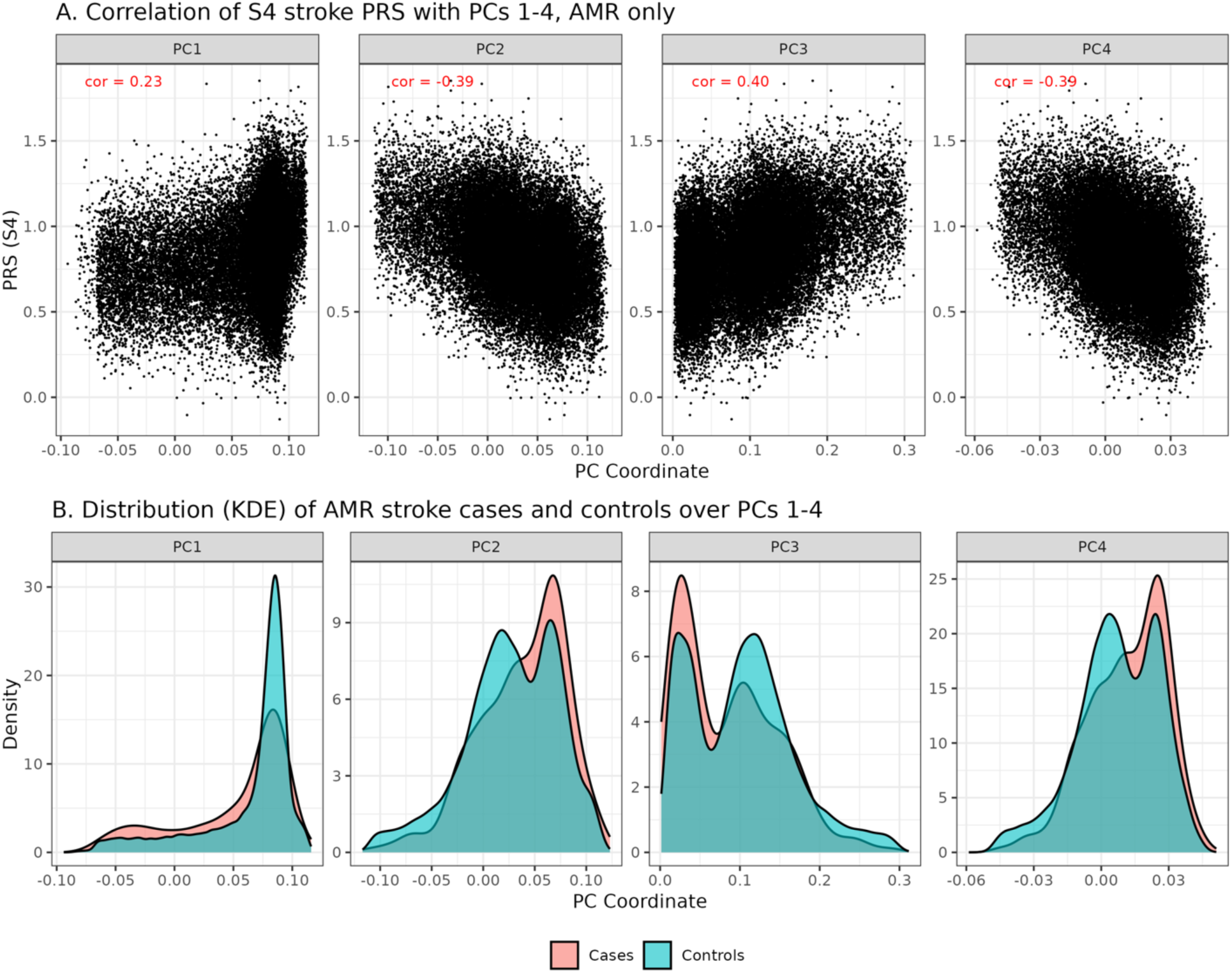
AMR stroke cases and controls differ in their distribution along PCs 1-4. In all four cases, kernel density estimates (B) show that more cases are found at PC levels that correlate with higher S4 PGS (A). LDpred2 produces similar results and is not plotted. Pearson correlation between the PGS and each principal component (cor) is shown in red.

**Figure S6:**
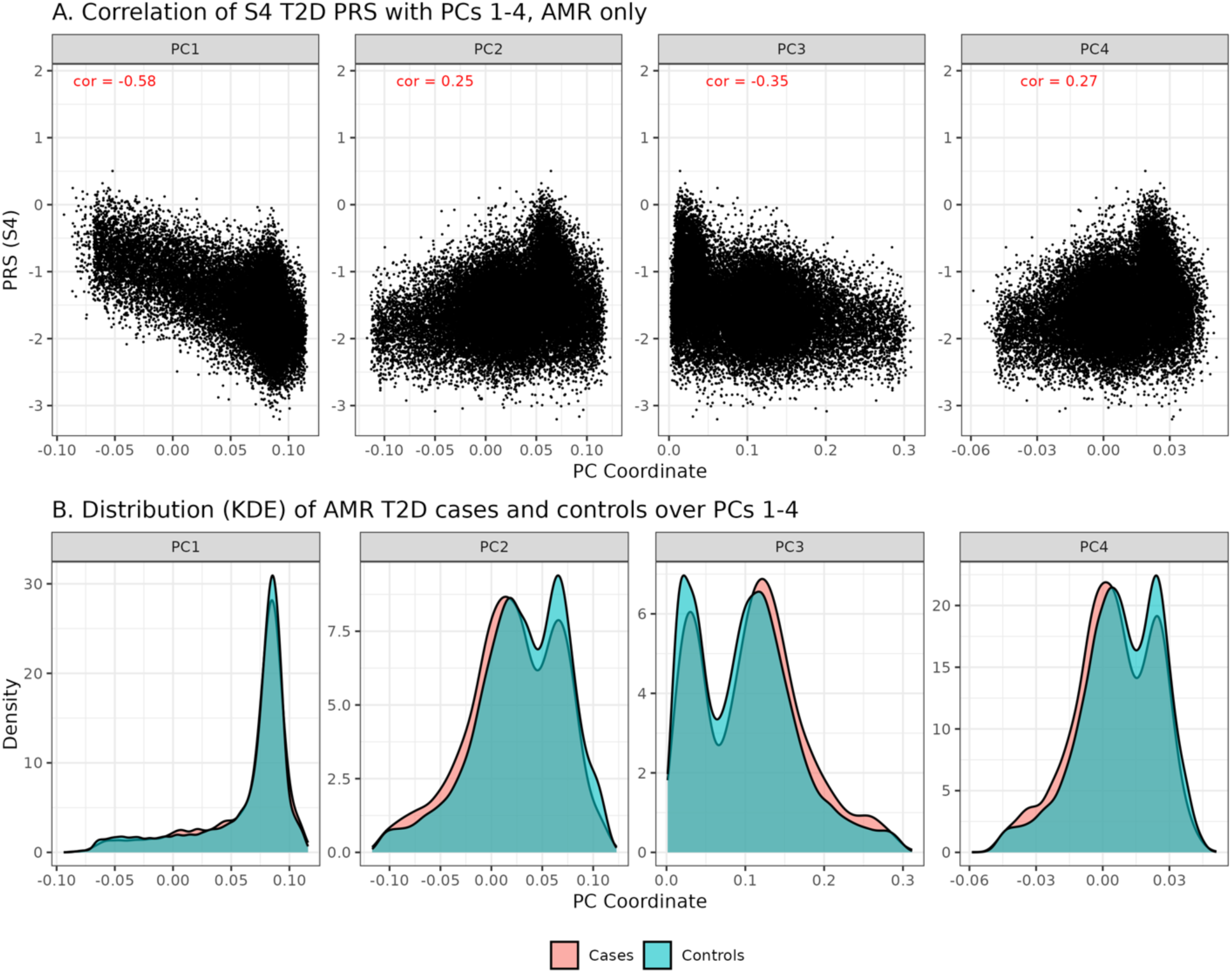
AMR type-2 diabetes (T2D) cases and controls also differ in their distributions along PCs 1-4 though to a lesser degree than stroke cases and controls. PCs 1-4 are weakly correlated with the S4 PGS (A). Similarly to stroke, population structure differences between cases and controls (B) push the PGSs in controls higher on average. Pearson correlation between the PGS and each principal component (cor) is shown in red.

**Table S1:** Breakdown of the 60 simulation runs per ancestry consisting of all 5 × 3 × 4 = 60 combinations of the three factors below

|  |  |
| --- | --- |
| **Genetic architectures:** | Fixed common SNP heritability with strong negative selection |
|  | Fixed per-SNP heritability with strong negative selection |
| | Fixed per-SNP heritability with strong negative selection ($r = 0.6$) |
|  | Fixed common SNP heritability with no negative selection |
|  | Fixed common SNP heritability with mild negative selection |
| **Causal SNP proportions:** | 0.01 |
|  | 0.001 |
| | $5 \times 10^{-4}$ |
| **Training sample sizes:** | 15,000 |
|  | 45,000 |
|  | 80,000 |
|  | 100,000 |

**Table S2:**
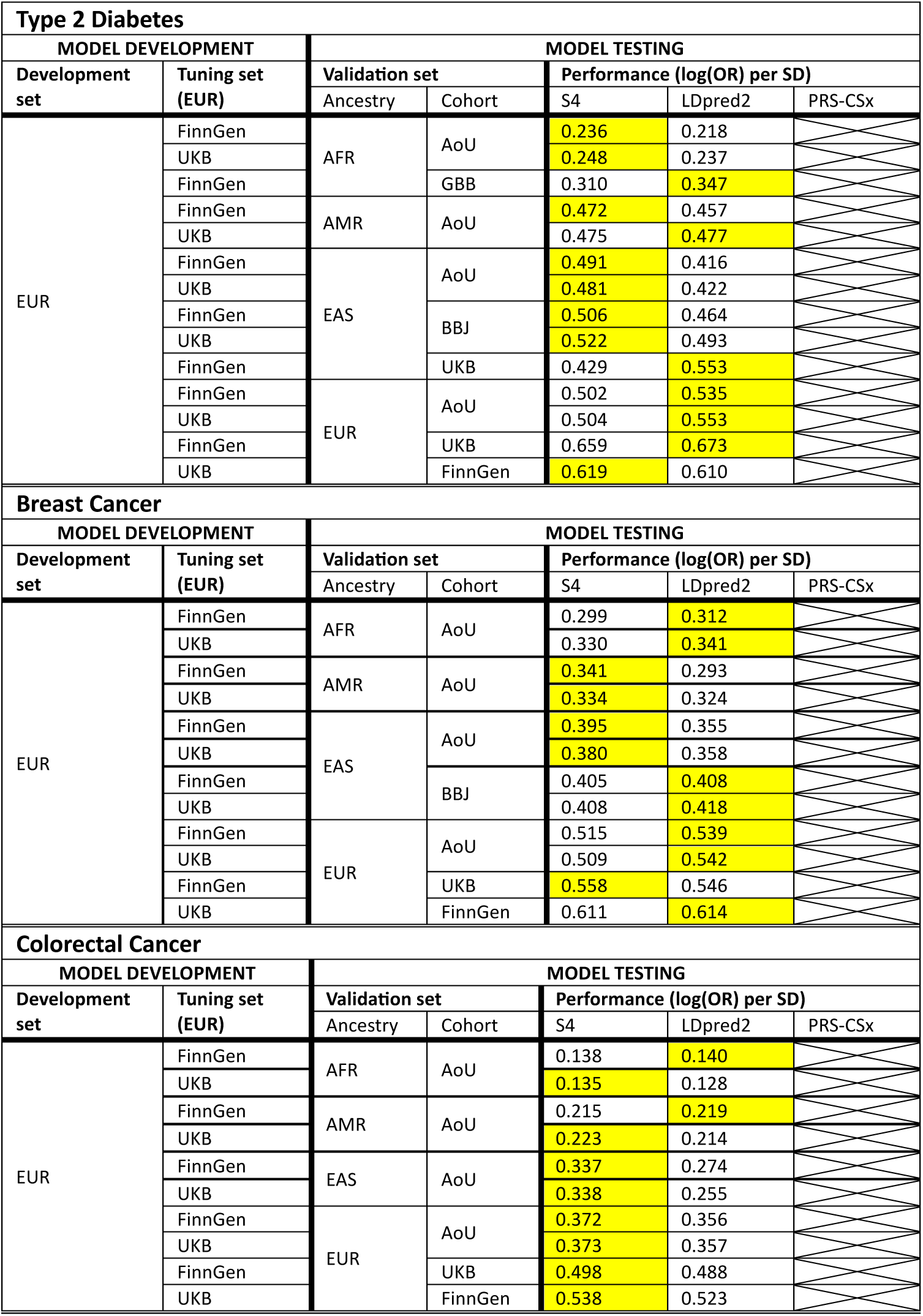
Performance of models trained on EUR-only data in log(OR) per SD, following the same format as Table 3 in the main text. The top performing method in each row in highlighted in yellow. Note that an EUR-only version of PRS-CSx was not fit, nor were EUR-only models for asthma and stroke.

**Table S3:**
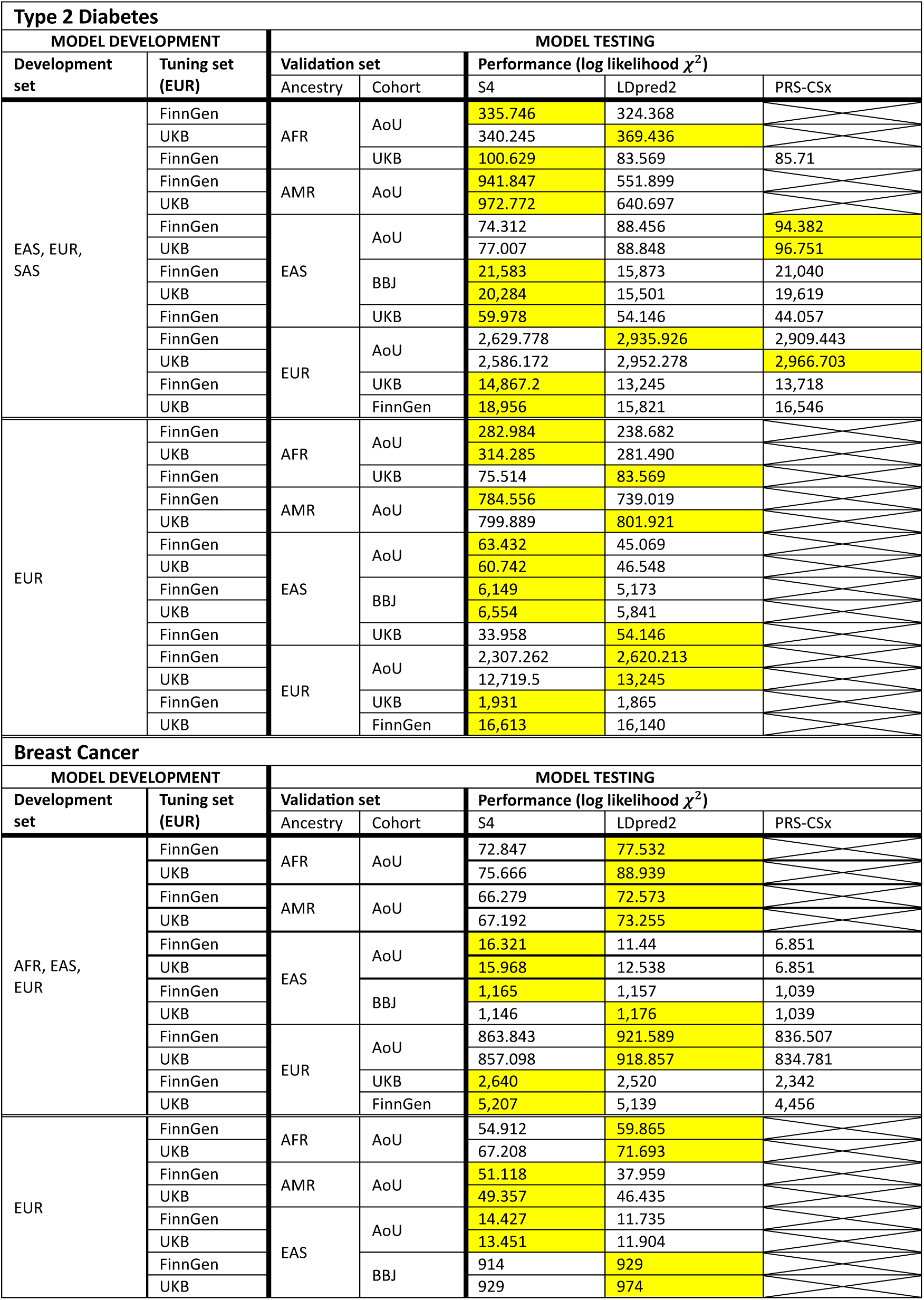
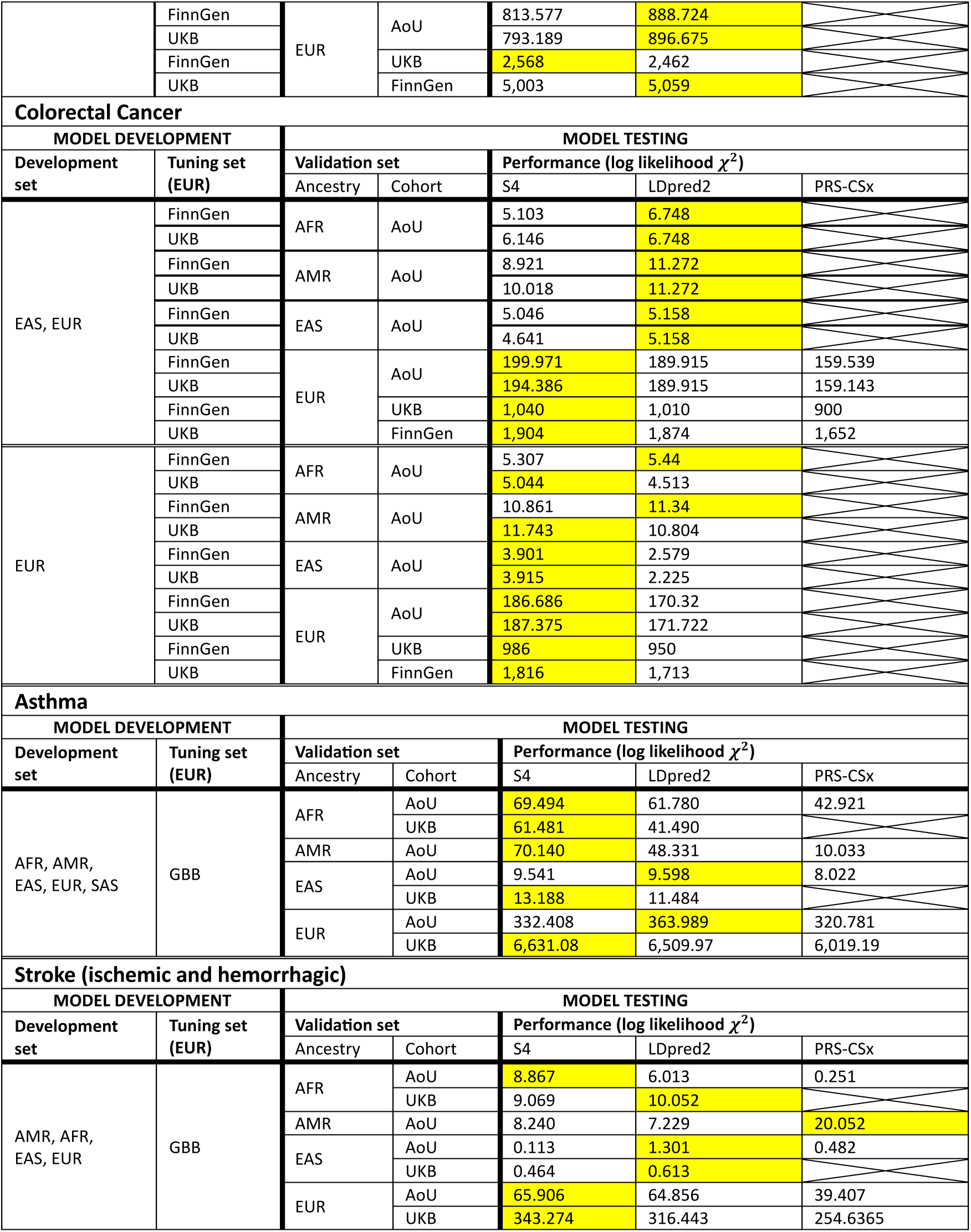
All indicated ancestry data sets used in model development consist of a consortium of available data from UK Biobank (UKB), FinnGen, and Biobank Japan (BBJ).

**Table S4i:**
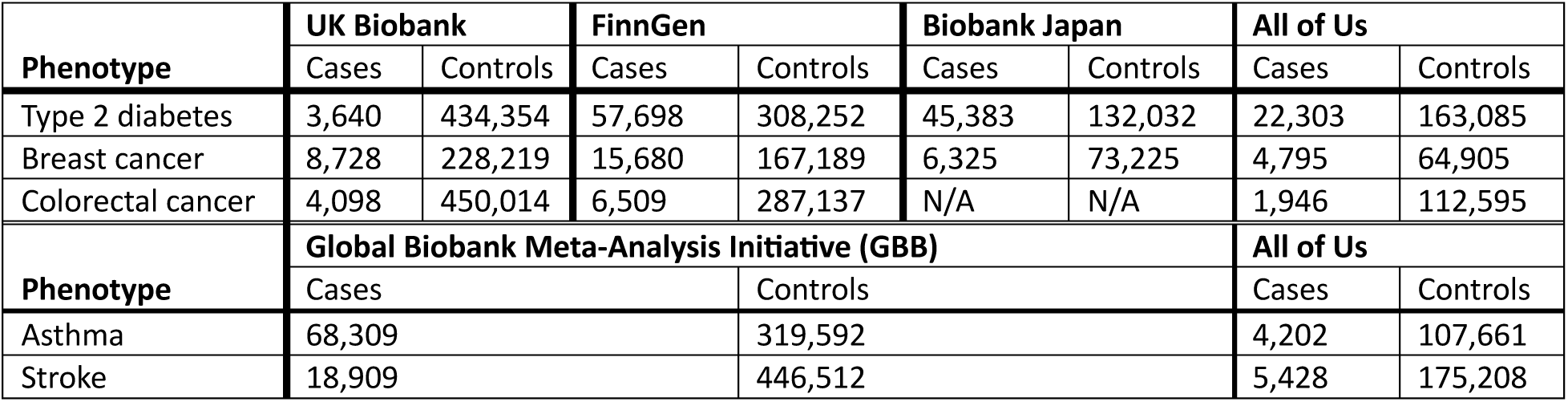
Case counts used for model validation in biobank tests by data source

| Phenotype | UK Biobank |  | FinnGen |  | Biobank Japan |  | All of Us |  |
| --- | --- | --- | --- | --- | --- | --- | --- | --- |
|  | Cases | Controls | Cases | Controls | Cases | Controls | Cases | Controls |
| Type 2 diabetes | 3,640 | 434,354 | 57,698 | 308,252 | 45,383 | 132,032 | 22,303 | 163,085 |
| Breast cancer | 8,728 | 228,219 | 15,680 | 167,189 | 6,325 | 73,225 | 4,795 | 64,905 |
| Colorectal cancer | 4,098 | 450,014 | 6,509 | 287,137 | N/A | N/A | 1,946 | 112,595 |
| Phenotype | Global Biobank Meta-Analysis Initiative (GBB) |  |  |  |  |  | All of Us |  |
|  | Cases |  |  | Controls |  |  | Cases | Controls |
| Asthma | 68,309 |  |  | 319,592 |  |  | 4,202 | 107,661 |
| Stroke | 18,909 |  |  | 446,512 |  |  | 5,428 | 175,208 |

**Table S4ii:** Case counts used for model validation in biobank tests stratified by predicted ancestry

| Phenotype | AFR |  | AMR |  | EAS |  | EUR |  |
| --- | --- | --- | --- | --- | --- | --- | --- | --- |
|  | Cases | Controls | Cases | Controls | Cases | Controls | Cases | Controls |
| Type 2 diabetes | 7,678 | 48,896 | 4,461 | 32,610 | 520 | 6,928 | 45,327 | 515,850 |
| Breast cancer | 749 | 19,386 | 460 | 14,365 | 141 | 3,102 | 12,281 | 256,379 |
| Colorectal cancer | 323 | 37,324 | 238 | 22,532 | 54 | 4,914 | 5,485 | 497,910 |
| Asthma | 2,285 | 31,803 | 482 | 22,361 | 309 | 4,612 | 69,435 | 368,477 |
| Stroke | 3,819 | 47,878 | 667 | 31,140 | 100 | 6,152 | 21,087 | 536,550 |

**Table S5:** Both S4 and LDpred2 PGSs are weakly correlated (Pearson correlation between 0.20 and 0.40 in magnitude) with the first four principal components (PCs) in AMR target populations. In no other target ancestry is the correlation between PCs 1-5 and either PGS that exceeds 0.14 in magnitude.

| Ancestry | PC1 correlation |  | PC2 correlation |  | PC3 correlation |  | PC4 correlation |  | PC5 correlation |  |
| --- | --- | --- | --- | --- | --- | --- | --- | --- | --- | --- |
|  | S4 | LDpred2 | S4 | LDpred2 | S4 | LDpred2 | S4 | LDpred2 | S4 | LDpred2 |
| AFR | -0.03 | -0.07 | -0.04 | -0.07 | 0.03 | 0.02 | -0.03 | -0.06 | 0.02 | 0.04 |
| AMR | 0.23 | 0.21 | -0.39 | -0.39 | 0.40 | 0.39 | -0.39 | -0.38 | 0.08 | 0.08 |
| EAS | -0.09 | -0.04 | 0.13 | 0.06 | -0.05 | 0.00 | -0.14 | -0.07 | -0.05 | -0.03 |
| EUR | 0.04 | 0.06 | 0.02 | 0.05 | 0.06 | 0.09 | 0.06 | 0.10 | 0.05 | 0.08 |

**Tables S6i-v:** Number of variants in each PGM in biobank tests* Details on hyperparameter tuning and model fitting are available in the Supplementary Materials.

| Development set | Number of variants in PGM |  |  |
| --- | --- | --- | --- |
|  | S4 | LDpred2 | PRS-CSx |
| AFR, EUR, SAS, EAS | 987,878 | 1,213,263 | 1,155,259 |
| EUR | 789,795 | 1,232,919 |  |

**ii. Breast Cancer:**
| Development set | Number of variants in PGM |  |  |
| --- | --- | --- | --- |
|  | S4 | LDpred2 | PRS-CSx |
| AFR, EUR, SAS, EAS | 390,110 | 1,327,214 | 1,295,848 |
| EUR | 381,748 | 1,335,689 |  |

**iii. Colorectal Cancer:**
| Development set | Number of variants in PGM |  |  |
| --- | --- | --- | --- |
|  | S4 | LDpred2 | PRS-CSx |
| AFR, EUR, SAS, EAS | 285,286 | 1,238,056 | 1,079,695 |
| EUR | 288,837 | 1,248,371 |  |

**iv. Asthma:**
| Development set | Number of variants in PGM |  |  |
| --- | --- | --- | --- |
|  | S4 | LDpred2 | PRS-CSx |
| AFR, EUR, SAS, EAS | 1,260,953 | 1,382,751 |  |

**v. Stroke:**
| Development set | Number of variants in PGM |  |  |
| --- | --- | --- | --- |
|  | S4 | LDpred2 | PRS-CSx |
| AFR, EUR, SAS, EAS | 959,427 | 1,298,127 |  |

## Footnotes

* Details on hyperparameter tuning and model fi4ng are available in the Supplementary Materials.

